# The clinical and epidemiological features and hints of 82 confirmed COVID-19 pediatric cases aged 0-16 in Wuhan, China

**DOI:** 10.1101/2020.03.15.20036319

**Authors:** Hui Yu, Qinzhen Cai, Xiang Dai, Xiuzhen Liu, Hong Sun

**Affiliations:** Wuhan Children’s Hospital, Tongji Medical School, Huazhong University of Science and Technology, Wuhan, China

## Abstract

Although COVID-19 pediatric patients just account for 1% of the overall cases, they are nonnegligible invisible infection sources. We quantitatively analyzed the clinical and epidemiological features of 82 confirmed cases aged 0-16 admitted to Wuhan Children’s Hospital, which are expected to shed some lights onto the pediatric diagnosis and therapy.

## Text

The outbreak of COVID-19 epidemic has attracted worldwide attentions since Dec., 2019, especially in Wuhan. The confirmed children just account for 1% of the overall confirmed cases. Although most of them have mild cases, they are still invisible infection sources of COVID-19 apt to be ignored. This study was carried out on Feb. 1-20, 2020 at Wuhan Children’s Hospital, the sole designated hospital for COVID-19 children patients. The participants consisted of 813 children receiving COVID-19 nucleic acid detection. We obtained epidemiological, clinical laboratory, and outcome data. The study was approved by Wuhan Children’s Hospital Ethics Committee, and official informed consent was obtained from guardians involved before enrollment when data were collected retrospectively.

Take throat-swab, anal-swab or urine specimens at admission, and implement real-time RT-PCR (Wuhan Huada Medical Technology Co., Ltd.). We described symptoms on admission; laboratory results; chest radiography and CT findings; treatment received for COVID-19 and clinical outcomes. Totally 812 children received COVID-19 nucleic acid detection. Therein, 82 (10.99 %) were infected with COVID-19. 74 (90.24%) had a history of exposure to confirmed or suspected family members. All the children lived in Wuhan.

As shown in Table 1, 51 boys and 31 girls were infected with COVID-19 with minimal and maximal ages being 3 days after birth and 16 years, respectively. Male infection rate (62.20%) is higher than female (37.8%), demonstrating the opposite to [1]. Children over 6-age have the highest infection rate (42.68%). On admission, most had fever or cough. Most have mild clinical symptoms with 8 no symptoms. 8 critically ill cases was found with 4 children and 4 infants. On admission, leucocytes were above (resp. below) normal in 21 (25.61%) patients (resp. 4 (4.88%)). The percentage of lymphocytes were above (resp. below) normal range in 48 (58.54%) patients (resp. 16 (19.51%)). Activated partial thromboplastin time (APTT) were above (resp. below) normal range in 9 (9/49, 18.37%) patients (resp. 3 (3/49, 6.12%)). Albumin were below normal range in 30(36.59%). γGlutamyl transpeptidase were above normal range in 27(32.93%). 30 (36.59%) had myocardial damage. Elevation of lactate dehydrogenase (LDH) in 15 (18.29%).

**Table 1:**
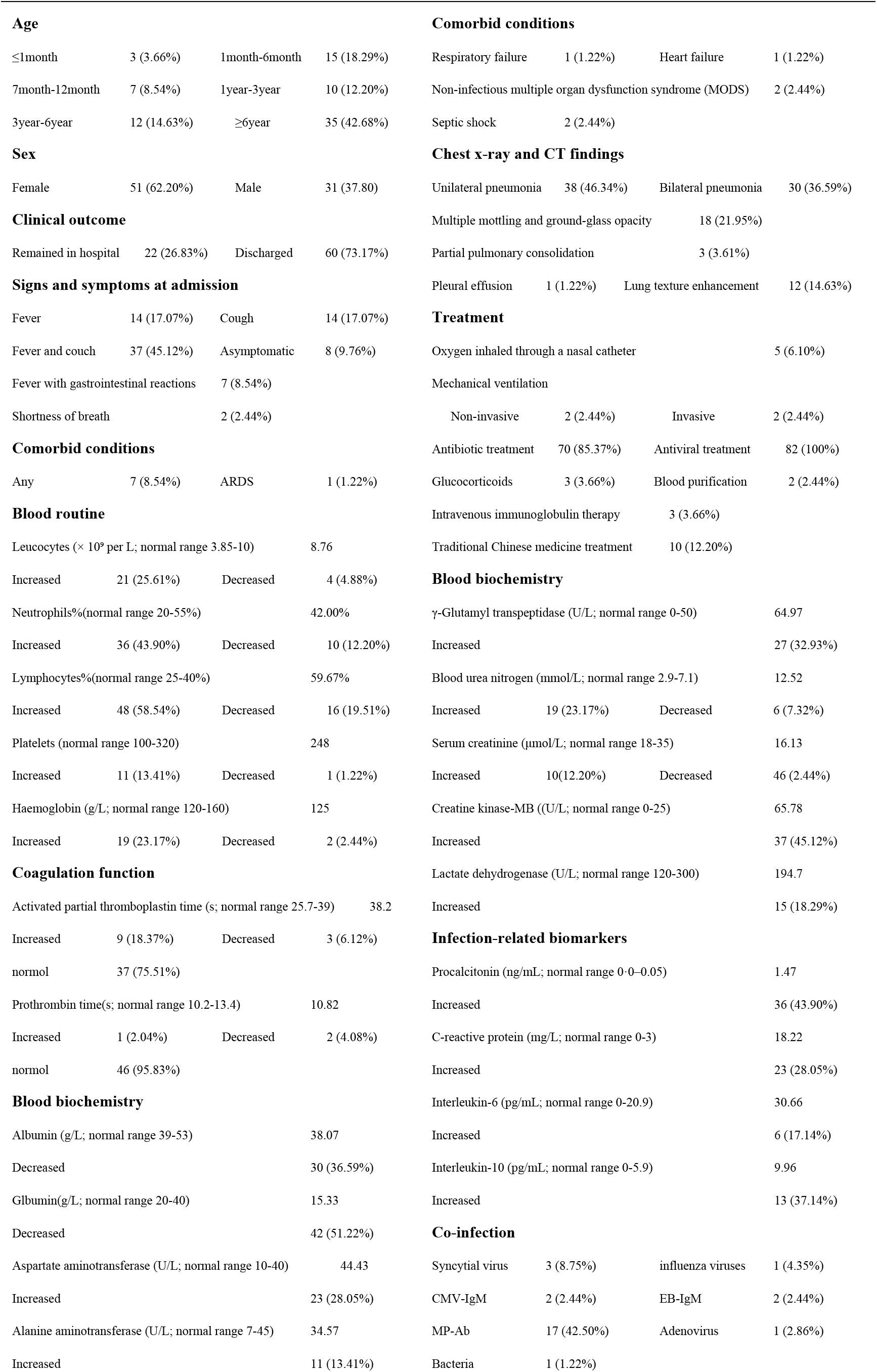
Clinical /laboratory features and treatment of 82 pediatric COVID-19 cases, in Wuhan.

Regarding chest X-ray and CT, 30 (36.59%), 38 (46.34%) and 18 (21.95%) patients showed bilateral, unilateral, multiple mottling and groundglass opacity pneumonia, respectively. 1 (1.22%) was found pleural effusion occurred. 2 (2.44%) had normal chest CTs.

All patients were treated in isolation and interferon atomization therapy. Most were given antibiotic or antiviral treatment. 6 (6.10%) children received nasal catheter oxygen therapy. Only 8 was transferred to ICU and given intensive care. 3 had serious complications. Till Feb. 20, 60 (73.17%) patients had been discharged whereas others still in hospital. Two discharged. The mean hospital stay was 11.2 day.

From the clinical and treatment data collected in 82 hospitalized COVID-19 patients under 16-age, it surfaces that the juvenile case size is much smaller than the adult counterparts [2,3] due to milder symptoms. Accordingly, they are apt to be ignored as potential infection resources of COVID-19. The laboratory results and clue are highlighted below, which might be beneficial for diagnosis and therapy for pediatric patients.

Most patients had a history of exposure to COVID-19 pneumonia confirmed or suspected family members, and children over 6-age have the highest infection rate. Opposite to [1], male proportion is higher than female.

Most have fever or cough on admission. Most have mild symptoms.

Discharged rate is 73.17%. Mean hospital stay was 11.2 days. Zero death rate.

Among inspected clinical features, the most important indexes for COVID-19 are

CT pneumonia, lymphocytes, APTT, Albumin, γGlutamyl transpeptidase, LDH.

The study was limited to a small number of patients from a single center in Wuhan. Further studies from multiple centers on a larger cohort would be beneficial to further validate the proposed route as well as understand of the disease.

## Data Availability

We claim the availability of the data referred in the manuscript was approved by Wuhan Children’s Hospital Ethics Committee, and official informed consent was obtained from guardians involved before enrolment when data were collected retrospectively.

## Notes

### Competing Interest Statement

The authors have declared no competing interest.

### Funding Statement

This research is supported by Wuhan Scientific and Technology Bureau Project with Grant No. 2017060201010161.

